# Human papillomavirus (HPV) infection and vaccination: a cross-sectional study of college students’ knowledge, awareness, and attitudes in Villanova, PA

**DOI:** 10.1101/2020.12.21.20248626

**Authors:** Jennifer A. Goldfarb, Joseph D. Comber

## Abstract

Human papillomaviruses are major causative agents of multiple cancers including cervical, penial, and oropharyngeal cancers. Almost all sexually active individuals are exposed to HPV in their lifetime and although not all HPV subtypes are capable of causing cancers, several high-risk subtypes widely circulate. Several HPV vaccines have been developed and successfully utilized to limit the spread of these viruses and reduce rates of associated cancers. Despite their success, HPV vaccination rates remain low. Studies estimate the highest prevalence of HPV in the United States is among college students. This makes college students an ideal target for interventions that promote HPV vaccination and prevention. To this end, we were interested in investigating the relationship between low HPV vaccine uptake and attitudes and awareness about HPV vaccination among college aged students. We designed a survey to assess knowledge and perception of HPV and HPV vaccination that could help pinpoint correlations between this knowledge and vaccination status. We find that more robust education focusing on the benefits of HPV vaccination could be utilized to improve vaccination rates.

## Introduction

Human papillomaviruses are non-enveloped double stranded DNA viruses in the *Papillomaviridae* family. Over 170 human papillomaviruses are known and at least 40 are capable of infecting the reproductive tract (among other tissues). Although many of these viruses establish an infection that is cleared by the host immune system, a small subset of viruses is capable of establishing chronic infections. High risk types of HPV are the most concerning since these are linked to cancer development, causing upwards of five percent of all cancers globally^1^. HPV is the leading cause of cervical cancer and causes most anal cancers (90%), oropharyngeal cancers (70%), and penile cancers (60%). In total, HPV is estimated to cause 570,000 cases of cancer in women and 60,000 cases of cancer in men worldwide every year.^2^

Vaccination is a highly effective intervention that protects against HPV infections, and therefore prevents the cancers that these viruses cause. Three vaccines against high-risk HPV types have been licensed in the United States, although only one of these vaccines is currently in use. All three HPV vaccines utilize virus-like particle technology, with the major capsid protein L1 assembling into a particle resembling HPV virions. Importantly, these VLPs do not contain viral DNA and do not have the ability to establish infection or cause cancer. Two of the earliest HPV vaccines were Cervarix, a bivalent vaccine developed by GSK protecting against HPV 16 and 18, and Gardasil, a quadrivalent vaccine developed by Merck protecting against HPV 6, 11, 16, and 18. Although these vaccines were successful in preventing infections^3^, they are no longer the preferred HPV vaccine and therefore are no longer distributed in the United States. The current preferred vaccine is Gardasil-9, a nonvalent vaccine developed by Merck that protects against nine distinct HPV types (6, 11, 16, 18, 31, 33, 45, 52, and 58). This vaccine is estimated to prevent 32,000 of the 35,000 cancers caused by HPV per year.^4^

Human papillomaviruses (HPV) are the most common sexually transmitted infections in the United States, and nearly all sexually active men and women will be infected by some type of HPV in their lifetime.^5^ Because vaccination is an important intervention to prevent infection, it is critical that individuals receive Gardasil 9 before becoming sexually active (recommended age 11-12). Studies demonstrate robust and durable immune responses in response to vaccination^6,7,8^ and some data suggest that the immune response of younger people is more robust than that of people in their late teens and early twenties^8^. Additionally, the CDC’s 2019 Youth Risk Behavior Survey (YRBS) reports that 27.4% of 9^th^ through 12^th^ grade students in the United States are currently sexually active and only 54.3% used a condom during their last sexual intercourse. Of these students, 3.0% reported being sexually active before 13 years old.^9^

Despite the importance of early vaccination against HPV, HPV vaccine uptake lags behind that of other vaccines recommended to be administered around the same age. For example, according to the National Immunization Survey (NIS-Teen, 2019), 90.2% of adolescents between 13-18 years of age were vaccinated against tetanus, diphtheria, and pertussis (Tdap) and 88.9% were vaccinated against meningococcus (MenACWY)^10^. In stark contrast, only 71.5% of adolescents in this age group were vaccinated with at least one dose of HPV vaccine and only 54.2% of adolescents were up to date on their HPV vaccine series.^10^ In Pennsylvania, the site of the current study, the numbers are similar with 77% of adolescents being vaccinated with at least one dose of Tdap and only 60.1% being up to date in the vaccine series.^10^

Studies estimate the highest prevalence of HPV in the United States to be among women visiting STD clinics and college students.^11^ This, combined with the lower vaccine uptake in adolescence, makes college students an ideal target for interventions that promote HPV vaccination and prevention. Surveys among college students similar to the one we conducted have occurred throughout the United States including in Florida^12^, Southern California^13^, South Carolina^14^, Michigan^15^, Mississippi^16^, Pennsylvania^17^, and Utah^18^. Similar studies have also been conducted outside of the United States in India^19,20^, Turkey^21^, and Vietnam^18^.

In an effort to boost HPV vaccination rates, it is necessary to investigate the relationship between low HPV vaccine uptake and the public’s attitudes and awareness about HPV vaccination. This is likely a multifaceted issue and there may be multiple reasons why HPV vaccination rates are lower than those of other vaccines, for example the Td/Tdap and MenACWY vaccination rates recommended for the same age group. Our survey focused on the college population to assess college students’ knowledge and awareness of HPV infection and HPV vaccination along with their HPV vaccination status. We sought to pinpoint possible correlations between knowledge, awareness, and/or perceptions of HPV infection and HPV vaccination that may be linked to vaccination status. The information we found in this study can be used to better educate individuals, dispel myths and stigmas, and increase awareness surrounding HPV infection and HPV vaccination. From this knowledge, we can also create initiatives to promote HPV vaccination not only among college students, but possibly among the recommended HPV vaccination age demographic (11-12) by considering what college students stated as obstacles or aversions to vaccination.

## Materials and methods

In order to better understand the knowledge, awareness, and attitudes of college-aged students towards the HPV vaccine, we conducted a cross-sectional study among university students at Villanova University (Villanova, PA) between February 13^th^ and March 9^th^, 2020. A convenience sampling method was used to recruit undergraduate and graduate students at least 18 years of age who were current students of Villanova at the time of survey completion. The survey was distributed through email, promotional flyers, and word-of-mouth and was hosted on a secure Google Forms account (G-Suite). All respondents provided informed consent before completing the survey and survey responses were collected in an anonymous manner. This study was approved by the Villanova University Institutional Review Board (IRB-FY2020-62).

The survey consisted of 32 multiple choice questions and 1 opened ended question ^Table 2^, similar to previous studies^12–21^. Questions asked participants about demographic information, general knowledge of HPV infection, perceived vulnerability of HPV infection, general knowledge of HPV vaccines, perceptions about HPV vaccine effectiveness and safety, and a question about HPV vaccination status. The open-ended question asked students to identify any information that could be helpful to convince them to get the vaccine.

Data was analyzed using chi-squared analyses to look for significant associations between: a) biological sex and response to survey questions Q6 through Q32; b) awareness of the existence of HPV and being vaccinated for HPV; c) perception of Gardasil 9 as safe and being vaccinated for HPV; d) approval by family/friends of Gardasil 9 vaccination and being vaccinated for HPV; e) correct response to knowledge section of the questionnaire (Q11 through Q21) and being vaccinated for HPV; f) perceived risk for HPV infection (no risk, low risk, moderate risk, high risk) and being vaccinated for HPV. P-values <0.05 were considered statistically significant in this study.

## Results

### General Demographic Information

**Table 1** summarizes the demographic characteristics of the study population. We collected responses from 217 participants (n=217). Forty-nine individuals reported as male (22.59%), 167 individuals reported as female (76.95%), and 1 individual preferred not to say (0.46%). Data from the respondent who preferred not to state biological sex was removed in an effort to maintain anonymity of individual responses. Participants ranged in age from 18 to 25+ with the majority of students being between 18 and 22 years old (88.89%). Since the undergraduate population of Villanova is much larger than the graduate population, it was not surprising that most respondents were undergraduate students (92.59% vs. 6.48% graduate students and 0.92% of students who preferred not to identify their year). Similarly, respondents were largely white and non-Hispanic (80.55%) which coincides with the full-time student enrollment demographics for Villanova University (White/Caucasian 75%).^22^ As expected since Villanova University is a Catholic institution, most participants identified as Christian (80.55%).

**Table 1:** Demographic characteristics of participants.

| Question Number | Demographic Variables | Total, n (%)<br>(n=216) | Female, n (%)<br>(n=167) | Male, n (%)<br>(n=49) |
| --- | --- | --- | --- | --- |
| Q1 | **Age** |  |  |  |
|  | 18 | 30 (13.9) | 22 (13.2) | 8 (16.3) |
|  | 19 | 49 (22.7) | 38 (22.8) | 11 (22.4) |
|  | 20 | 47 (21.8) | 38 (22.8) | 9 (18.4) |
|  | 21 | 47 (21.8) | 32 (19.2) | 15 (30.6) |
|  | 22 | 19 (8.8) | 16 (9.6) | 3 (6.1) |
|  | 23 | 3 (1.4) | 2 (1.2) | 1 (2.0) |
|  | 24 | 5 (2.3) | 3 (1.8) | 2 (4.1) |
|  | 25+ | 16 (7.4) | 16 (9.6) | 0 (0.0) |
| Q3 | **Ethnicity** |  |  |  |
|  | White, non-Hispanic | 174 (80.6) | 135 (80.8) | 39 (80.0) |
|  | Black, non-Hispanic | 5 (2.3) | 5 (3.0) | 0 (0.0) |
|  | Hispanic or Latino | 17 (7.9) | 13 (7.8) | 4 (8.2) |
|  | Other | 20 (9.3) | 14 (8.4) | 6 (12.2) |
| Q4 | **Religion** |  |  |  |
|  | Christian | 174 (80.6) | 135 (80.8) | 39 (79.6) |
|  | Jewish | 2 (0.9) | 1 (0.6) | 1 (2.0) |
|  | Not religious | 34 (15.7) | 26 (15.6) | 8 (16.3) |
|  | Other | 4 (1.9) | 3 (1.8) | 1 (2.0) |
|  | Prefer not to say | 2 (0.9) | 2 (1.2) | 0 (0.0) |
| Q5 | **Academic Year** |  |  |  |
|  | Freshman | 52 (24.1) | 36 (21.6) | 16 (32.7) |
|  | Sophomore | 52 (24.1) | 45 (26.9) | 7 (14.3) |
|  | Junior | 35 (16.2) | 25 (15.0) | 10 (20.4) |
|  | Senior | 61 (28.2) | 48 (28.7) | 13 (26.5) |
|  | Graduate student | 14 (6.5) | 11 (6.7) | 3 (6.1) |
|  | Prefer not to say | 2 (0.9) | 2 (1.2) | 0 (0.0) |

Q2 of the survey asked respondents to state their gender. This data is located in the column headings.

**Table 2:**
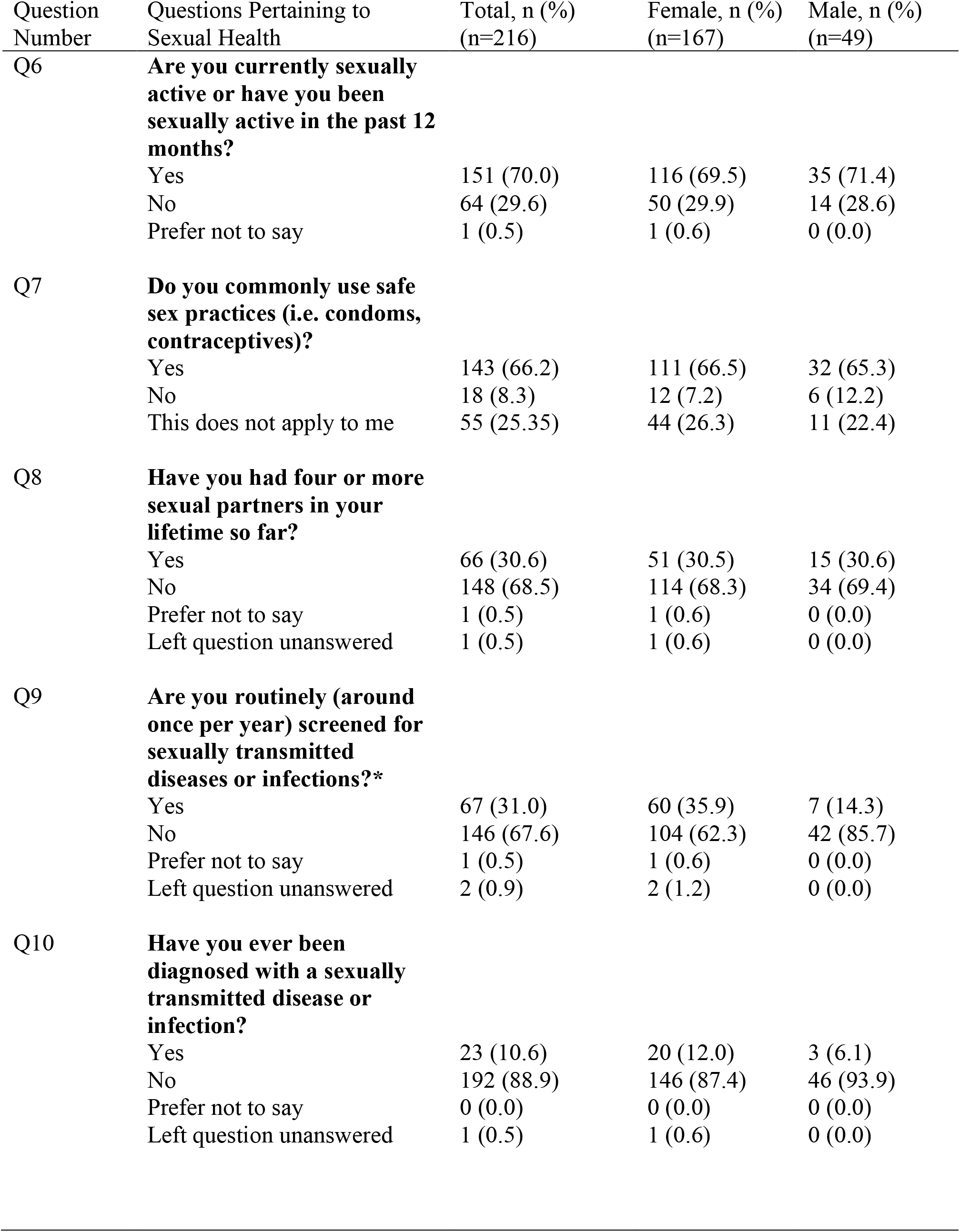
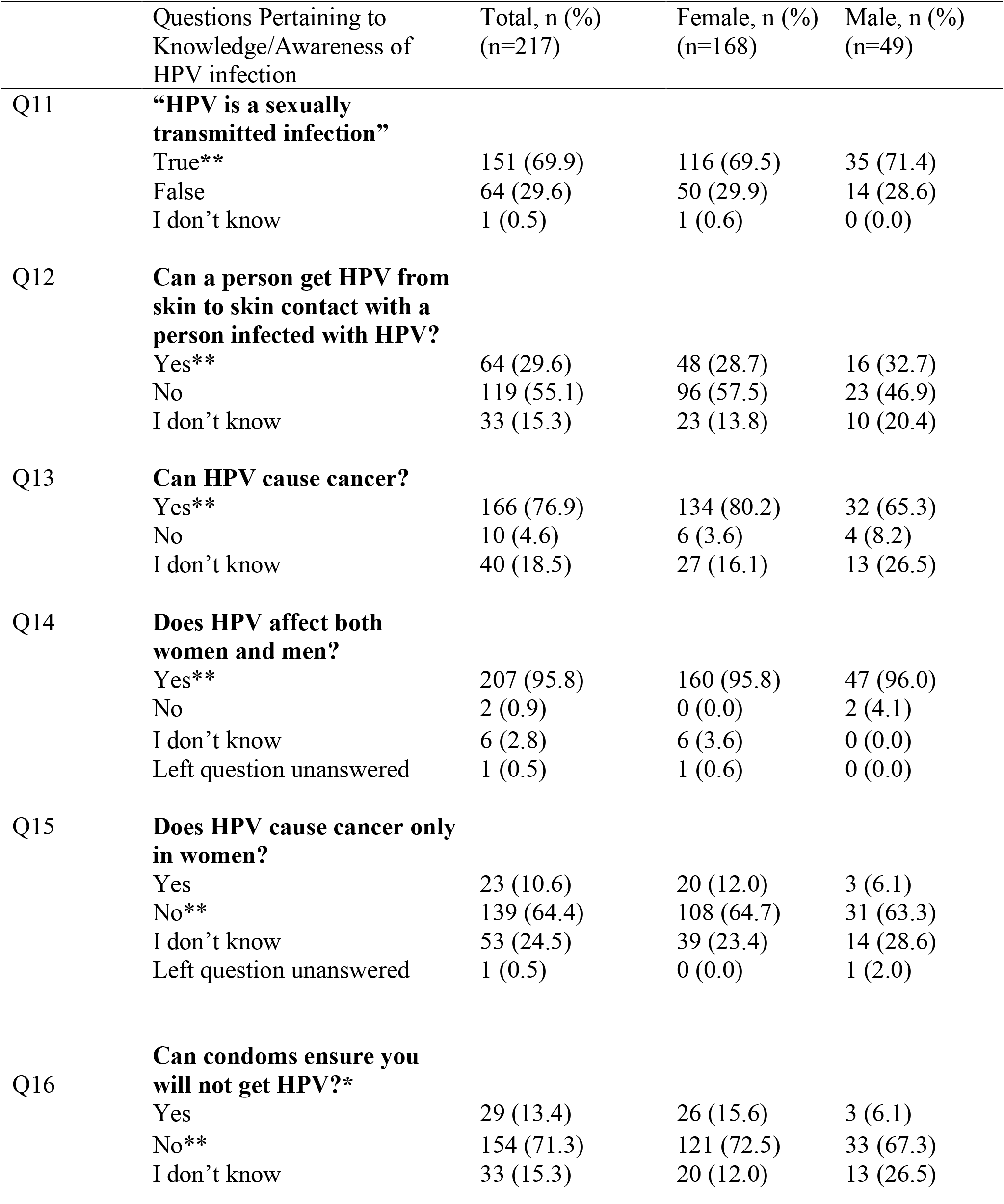
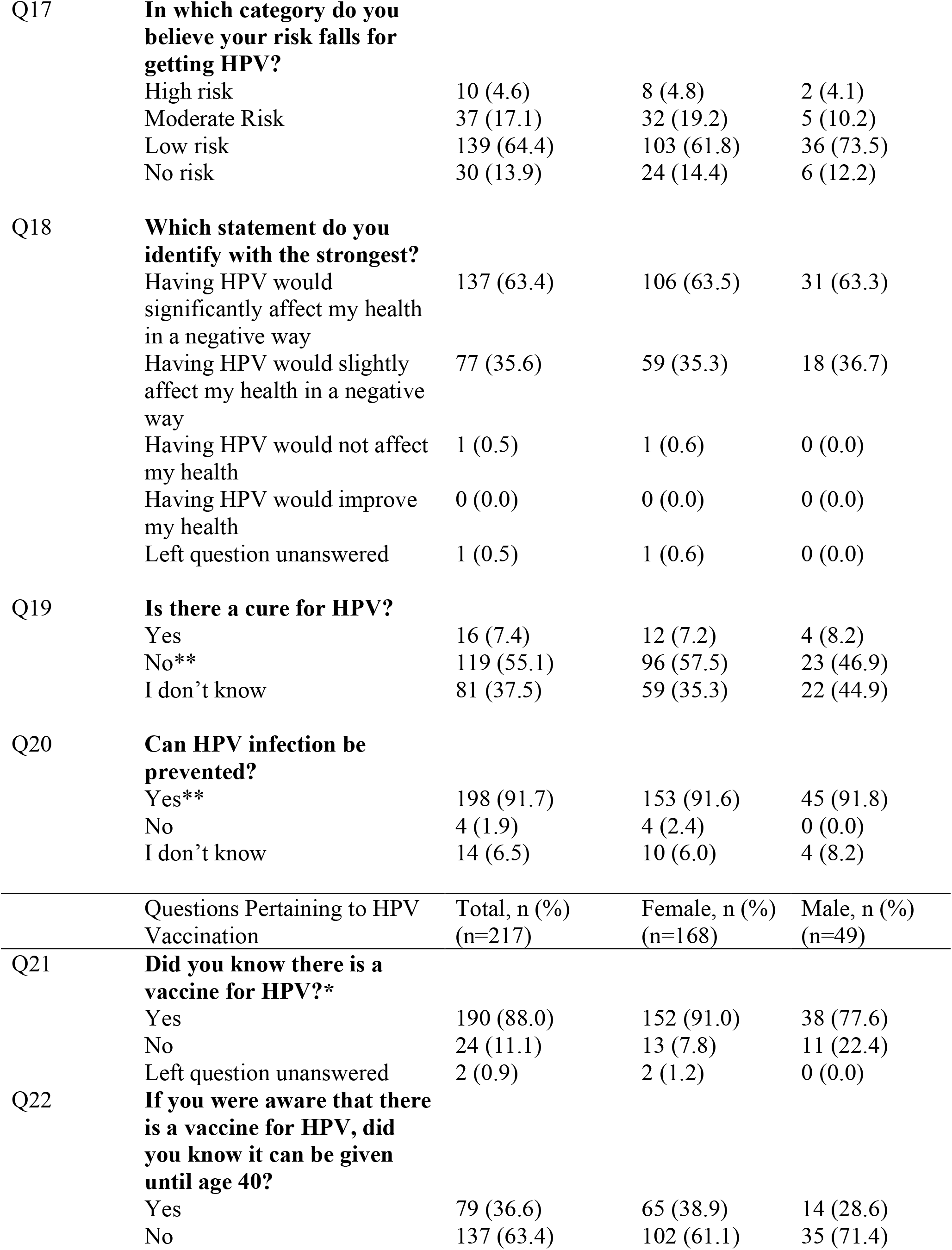
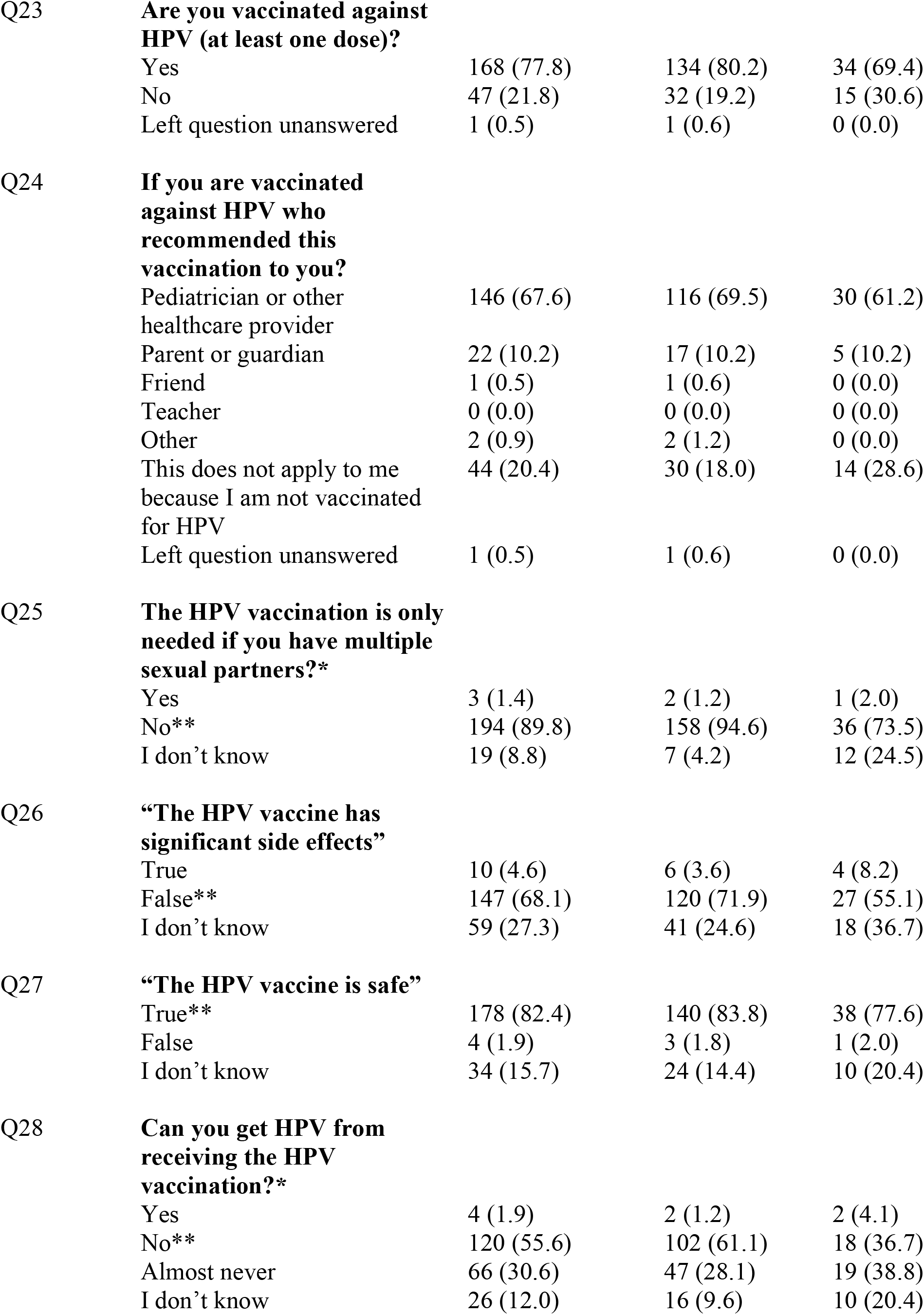
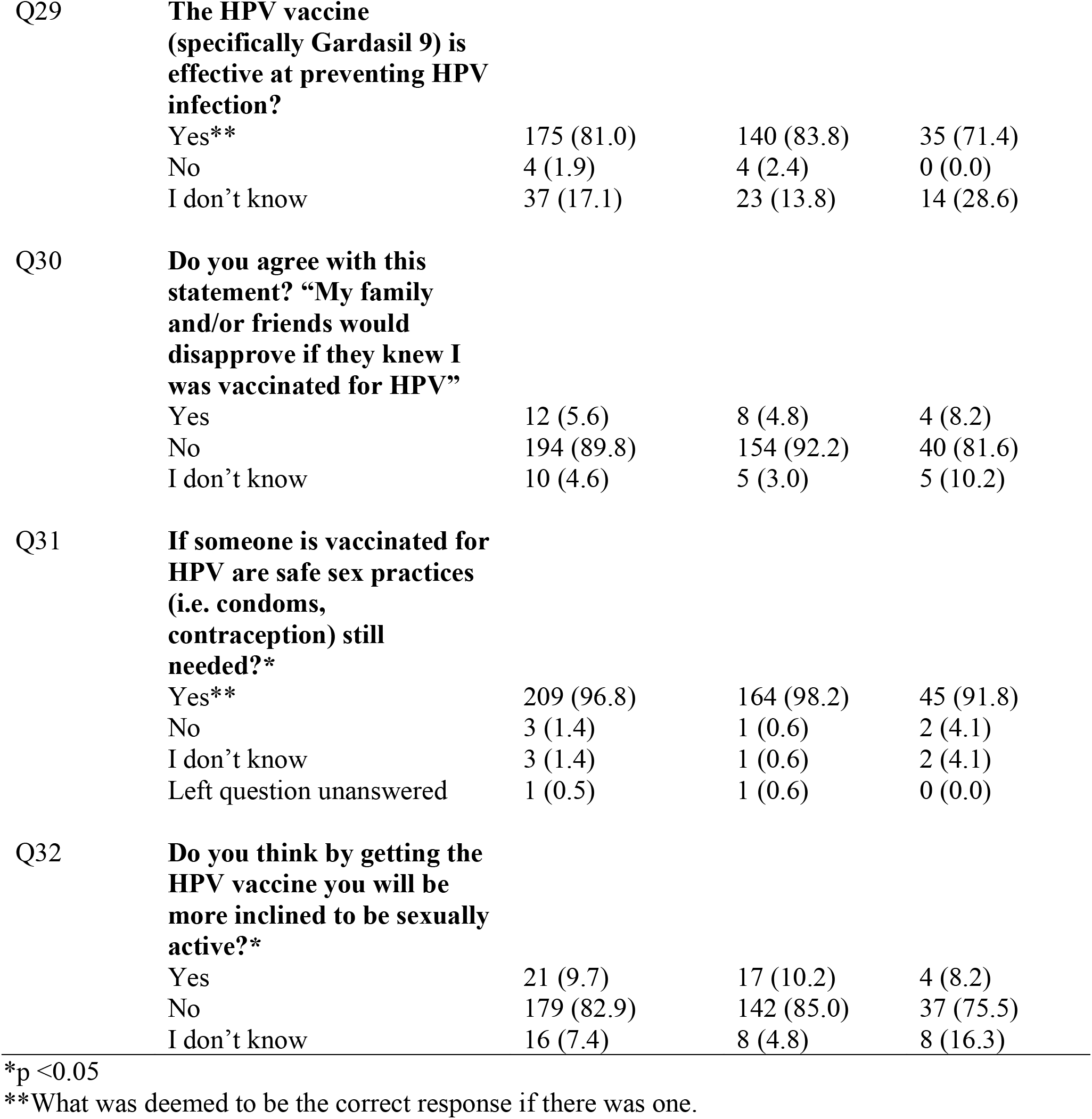
Total, female, and male responses to study questions.

| Question Number | Questions Pertaining to Sexual Health | Total, n (%)<br>(n=216) | Female, n (%)<br>(n=167) | Male, n (%)<br>(n=49) |
| --- | --- | --- | --- | --- |
| Q6 | **Are you currently sexually active or have you been sexually active in the past 12 months?** |  |  |  |
|  | Yes | 151 (70.0) | 116 (69.5) | 35 (71.4) |
|  | No | 64 (29.6) | 50 (29.9) | 14 (28.6) |
|  | Prefer not to say | 1 (0.5) | 1 (0.6) | 0 (0.0) |
| Q7 | **Do you commonly use safe sex practices (i.e. condoms, contraceptives)?** |  |  |  |
|  | Yes | 143 (66.2) | 111 (66.5) | 32 (65.3) |
|  | No | 18 (8.3) | 12 (7.2) | 6 (12.2) |
|  | This does not apply to me | 55 (25.35) | 44 (26.3) | 11 (22.4) |
| Q8 | **Have you had four or more sexual partners in your lifetime so far?** |  |  |  |
|  | Yes | 66 (30.6) | 51 (30.5) | 15 (30.6) |
|  | No | 148 (68.5) | 114 (68.3) | 34 (69.4) |
|  | Prefer not to say | 1 (0.5) | 1 (0.6) | 0 (0.0) |
|  | Left question unanswered | 1 (0.5) | 1 (0.6) | 0 (0.0) |
| Q9 | **Are you routinely (around once per year) screened for sexually transmitted diseases or infections?\*** |  |  |  |
|  | Yes | 67 (31.0) | 60 (35.9) | 7 (14.3) |
|  | No | 146 (67.6) | 104 (62.3) | 42 (85.7) |
|  | Prefer not to say | 1 (0.5) | 1 (0.6) | 0 (0.0) |
|  | Left question unanswered | 2 (0.9) | 2 (1.2) | 0 (0.0) |
| Q10 | **Have you ever been diagnosed with a sexually transmitted disease or infection?** |  |  |  |
|  | Yes | 23 (10.6) | 20 (12.0) | 3 (6.1) |
|  | No | 192 (88.9) | 146 (87.4) | 46 (93.9) |
|  | Prefer not to say | 0 (0.0) | 0 (0.0) | 0 (0.0) |
|  | Left question unanswered | 1 (0.5) | 1 (0.6) | 0 (0.0) |

|  | Questions Pertaining to Knowledge/Awareness of HPV infection | Total, n (%)<br>(n=217) | Female, n (%)<br>(n=168) | Male, n (%)<br>(n=49) |
| --- | --- | --- | --- | --- |
| Q11 | **“HPV is a sexually transmitted infection”** |  |  |  |
|  | True** | 151 (69.9) | 116 (69.5) | 35 (71.4) |
|  | False | 64 (29.6) | 50 (29.9) | 14 (28.6) |
|  | I don’t know | 1 (0.5) | 1 (0.6) | 0 (0.0) |
| Q12 | **Can a person get HPV from skin to skin contact with a person infected with HPV?** |  |  |  |
|  | Yes** | 64 (29.6) | 48 (28.7) | 16 (32.7) |
|  | No | 119 (55.1) | 96 (57.5) | 23 (46.9) |
|  | I don’t know | 33 (15.3) | 23 (13.8) | 10 (20.4) |
| Q13 | **Can HPV cause cancer?** |  |  |  |
|  | Yes** | 166 (76.9) | 134 (80.2) | 32 (65.3) |
|  | No | 10 (4.6) | 6 (3.6) | 4 (8.2) |
|  | I don’t know | 40 (18.5) | 27 (16.1) | 13 (26.5) |
| Q14 | **Does HPV affect both women and men?** |  |  |  |
|  | Yes** | 207 (95.8) | 160 (95.8) | 47 (96.0) |
|  | No | 2 (0.9) | 0 (0.0) | 2 (4.1) |
|  | I don’t know | 6 (2.8) | 6 (3.6) | 0 (0.0) |
|  | Left question unanswered | 1 (0.5) | 1 (0.6) | 0 (0.0) |
| Q15 | **Does HPV cause cancer only in women?** |  |  |  |
|  | Yes | 23 (10.6) | 20 (12.0) | 3 (6.1) |
|  | No** | 139 (64.4) | 108 (64.7) | 31 (63.3) |
|  | I don’t know | 53 (24.5) | 39 (23.4) | 14 (28.6) |
|  | Left question unanswered | 1 (0.5) | 0 (0.0) | 1 (2.0) |
| Q16 | **Can condoms ensure you will not get HPV?\*** |  |  |  |
|  | Yes | 29 (13.4) | 26 (15.6) | 3 (6.1) |
|  | No** | 154 (71.3) | 121 (72.5) | 33 (67.3) |
|  | I don’t know | 33 (15.3) | 20 (12.0) | 13 (26.5) |
| Q17 | **In which category do you believe your risk falls for getting HPV?** |  |  |  |
|  | High risk | 10 (4.6) | 8 (4.8) | 2 (4.1) |
|  | Moderate Risk | 37 (17.1) | 32 (19.2) | 5 (10.2) |
|  | Low risk | 139 (64.4) | 103 (61.8) | 36 (73.5) |
|  | No risk | 30 (13.9) | 24 (14.4) | 6 (12.2) |
| Q18 | **Which statement do you identify with the strongest?** |  |  |  |
|  | Having HPV would significantly affect my health in a negative way | 137 (63.4) | 106 (63.5) | 31 (63.3) |
|  | Having HPV would slightly affect my health in a negative way | 77 (35.6) | 59 (35.3) | 18 (36.7) |
|  | Having HPV would not affect my health | 1 (0.5) | 1 (0.6) | 0 (0.0) |
|  | Having HPV would improve my health | 0 (0.0) | 0 (0.0) | 0 (0.0) |
|  | Left question unanswered | 1 (0.5) | 1 (0.6) | 0 (0.0) |
| Q19 | **Is there a cure for HPV?** |  |  |  |
|  | Yes | 16 (7.4) | 12 (7.2) | 4 (8.2) |
|  | No** | 119 (55.1) | 96 (57.5) | 23 (46.9) |
|  | I don't know | 81 (37.5) | 59 (35.3) | 22 (44.9) |
| Q20 | **Can HPV infection be prevented?** |  |  |  |
|  | Yes** | 198 (91.7) | 153 (91.6) | 45 (91.8) |
|  | No | 4 (1.9) | 4 (2.4) | 0 (0.0) |
|  | I don't know | 14 (6.5) | 10 (6.0) | 4 (8.2) |
|  | Questions Pertaining to HPV Vaccination | Total, n (%)<br>(n=217) | Female, n (%)<br>(n=168) | Male, n (%)<br>(n=49) |
| Q21 | **Did you know there is a vaccine for HPV?\*** |  |  |  |
|  | Yes | 190 (88.0) | 152 (91.0) | 38 (77.6) |
|  | No | 24 (11.1) | 13 (7.8) | 11 (22.4) |
|  | Left question unanswered | 2 (0.9) | 2 (1.2) | 0 (0.0) |
| Q22 | **If you were aware that there is a vaccine for HPV, did you know it can be given until age 40?** |  |  |  |
|  | Yes | 79 (36.6) | 65 (38.9) | 14 (28.6) |
|  | No | 137 (63.4) | 102 (61.1) | 35 (71.4) |
| Q23 | **Are you vaccinated against HPV (at least one dose)?** |  |  |  |
|  | Yes | 168 (77.8) | 134 (80.2) | 34 (69.4) |
|  | No | 47 (21.8) | 32 (19.2) | 15 (30.6) |
|  | Left question unanswered | 1 (0.5) | 1 (0.6) | 0 (0.0) |
| Q24 | **If you are vaccinated against HPV who recommended this vaccination to you?** |  |  |  |
|  | Pediatrician or other healthcare provider | 146 (67.6) | 116 (69.5) | 30 (61.2) |
|  | Parent or guardian | 22 (10.2) | 17 (10.2) | 5 (10.2) |
|  | Friend | 1 (0.5) | 1 (0.6) | 0 (0.0) |
|  | Teacher | 0 (0.0) | 0 (0.0) | 0 (0.0) |
|  | Other | 2 (0.9) | 2 (1.2) | 0 (0.0) |
|  | This does not apply to me because I am not vaccinated for HPV | 44 (20.4) | 30 (18.0) | 14 (28.6) |
|  | Left question unanswered | 1 (0.5) | 1 (0.6) | 0 (0.0) |
| Q25 | **The HPV vaccination is only needed if you have multiple sexual partners?\*** |  |  |  |
|  | Yes | 3 (1.4) | 2 (1.2) | 1 (2.0) |
|  | No** | 194 (89.8) | 158 (94.6) | 36 (73.5) |
|  | I don't know | 19 (8.8) | 7 (4.2) | 12 (24.5) |
| Q26 | **“The HPV vaccine has significant side effects”** |  |  |  |
|  | True | 10 (4.6) | 6 (3.6) | 4 (8.2) |
|  | False** | 147 (68.1) | 120 (71.9) | 27 (55.1) |
|  | I don't know | 59 (27.3) | 41 (24.6) | 18 (36.7) |
| Q27 | **“The HPV vaccine is safe”** |  |  |  |
|  | True** | 178 (82.4) | 140 (83.8) | 38 (77.6) |
|  | False | 4 (1.9) | 3 (1.8) | 1 (2.0) |
|  | I don't know | 34 (15.7) | 24 (14.4) | 10 (20.4) |
| Q28 | **Can you get HPV from receiving the HPV vaccination?\*** |  |  |  |
|  | Yes | 4 (1.9) | 2 (1.2) | 2 (4.1) |
|  | No** | 120 (55.6) | 102 (61.1) | 18 (36.7) |
|  | Almost never | 66 (30.6) | 47 (28.1) | 19 (38.8) |
|  | I don't know | 26 (12.0) | 16 (9.6) | 10 (20.4) |
| Q29 | **The HPV vaccine (specifically Gardasil 9) is effective at preventing HPV infection?** |  |  |  |
|  | Yes** | 175 (81.0) | 140 (83.8) | 35 (71.4) |
|  | No | 4 (1.9) | 4 (2.4) | 0 (0.0) |
|  | I don't know | 37 (17.1) | 23 (13.8) | 14 (28.6) |
| Q30 | **Do you agree with this statement? "My family and/or friends would disapprove if they knew I was vaccinated for HPV"** |  |  |  |
|  | Yes | 12 (5.6) | 8 (4.8) | 4 (8.2) |
|  | No | 194 (89.8) | 154 (92.2) | 40 (81.6) |
|  | I don't know | 10 (4.6) | 5 (3.0) | 5 (10.2) |
| Q31 | **If someone is vaccinated for HPV are safe sex practices (i.e. condoms, contraception) still needed?\*** |  |  |  |
|  | Yes** | 209 (96.8) | 164 (98.2) | 45 (91.8) |
|  | No | 3 (1.4) | 1 (0.6) | 2 (4.1) |
|  | I don't know | 3 (1.4) | 1 (0.6) | 2 (4.1) |
|  | Left question unanswered | 1 (0.5) | 1 (0.6) | 0 (0.0) |
| Q32 | **Do you think by getting the HPV vaccine you will be more inclined to be sexually active?\*** |  |  |  |
|  | Yes | 21 (9.7) | 17 (10.2) | 4 (8.2) |
|  | No | 179 (82.9) | 142 (85.0) | 37 (75.5) |
|  | I don't know | 16 (7.4) | 8 (4.8) | 8 (16.3) |

\*p <0.05
\*\*What was deemed to be the correct response if there was one.

### Safe sex practices and routine screenings

**Figure 1** illustrates the respondents behavior surrounding safe sex and routine STD screening. At the time of the survey, 69.5% of females (n=116) and 71.4% of males (n=35) reported that they were currently sexually active or had been sexually active in the past 12 months. Additionally, 31.5% of females (n=51) and 30.6% of males (n=15) reported having four or more sexual partners in their lifetime. Although there was no significant statistical difference between females and males with respect to sexual history, number of partners, or history of sexually transmitted disease/infection (STD/STI), females were more likely than males to be routinely screened for STDs/STIs (p=0.003180, χ^2^=9.0470). A remarkably low number of participants in this survey indicated that they are routinely screened for these diseases (35.9% for females, 14.3% for males). Despite the recommendation that sexually active females under 25 get screened for STDs at least once per year^23^, only 48.0% of sexually active females under 25 (n=49) reported being routinely screened. The CDC also recommends annual STD testing for all sexually active gay and bisexual men^23^ and men in high prevalence clinical settings (adolescent clinics, correctional facilities, and STD clinics)^24^. Only 1.7% (n=6) of sexually active men reported being routinely screened for STDs. Since this data was collected in the university population, the surveyed males were not routinely in an identified high prevalence clinical setting that would warrant routine testing, but it cannot be determined from the data collected if the sexually active surveyed males are men who have sex with men.

**Figure 1:**
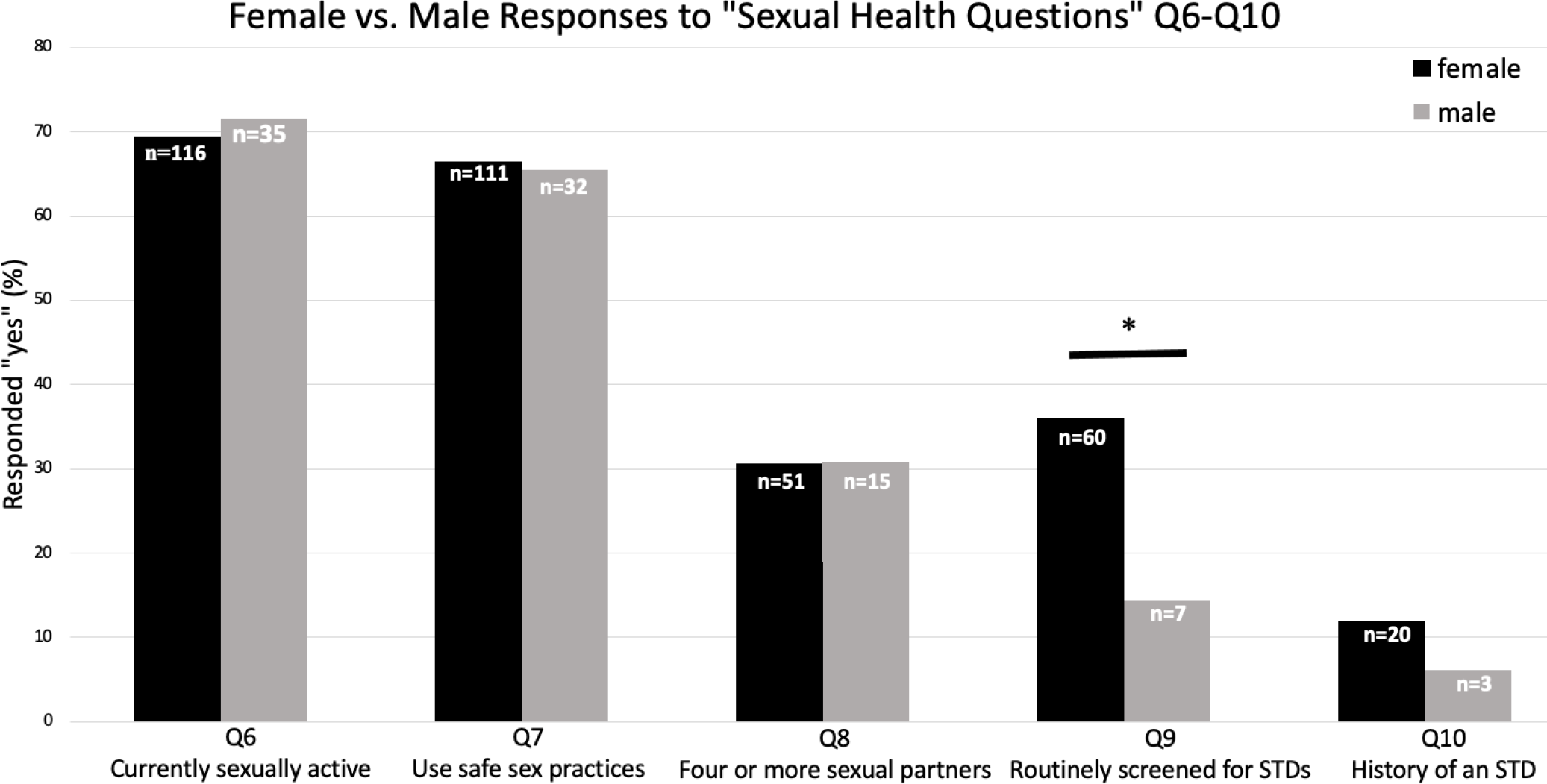
Females are more likely than males to be routinely screened for STDs/STIs. However, no other significant differences exist between males and females with respect to sexual health. *\*p<0.05*

### Knowledge about HPV infection, vaccination, and vaccine safety

The choice to be vaccinated is multifaceted, and factors that are thought to play a role include knowledge about the vaccine and disease, health care provider recommendations, perceptions about the risks of diseases, perceptions about vaccination effectiveness, and socioeconomic status^25,26,27^. We were interested in understanding more about these factors as they specifically relate to HPV vaccination status among college students. Therefore, we designed our survey to assess several potential factors that may drive these decisions. One such factor was knowledge about HPV infection itself, and to this end participants were surveyed on their knowledge and awareness about HPV infection (Q11-16, Q19-20, **Figure 2** and **Table 2)** The majority of participants correctly identified that HPV affects both male and females (95.8%), is a sexually transmitted disease (69.9%), and is capable of causing cancer (76.9%). However, a much lower percentage of participants understood that HPV can be spread by skin to skin contact with only 29.6% of participants responding correctly to this question (55.1% incorrectly) and 15.3% of participants unsure. Interestingly, participants (both male and female) who responded correctly to all eight knowledge questions were not significantly more likely to be vaccinated for HPV (with at least one dose) (p= 0.1196, χ^2^=4.1010). Overall, this data suggest that factors beyond knowledge of the infectious disease play more significant roles in the decision to vaccinate in line with what has previously been reported for HPV vaccination.^26^

**Figure 2:**
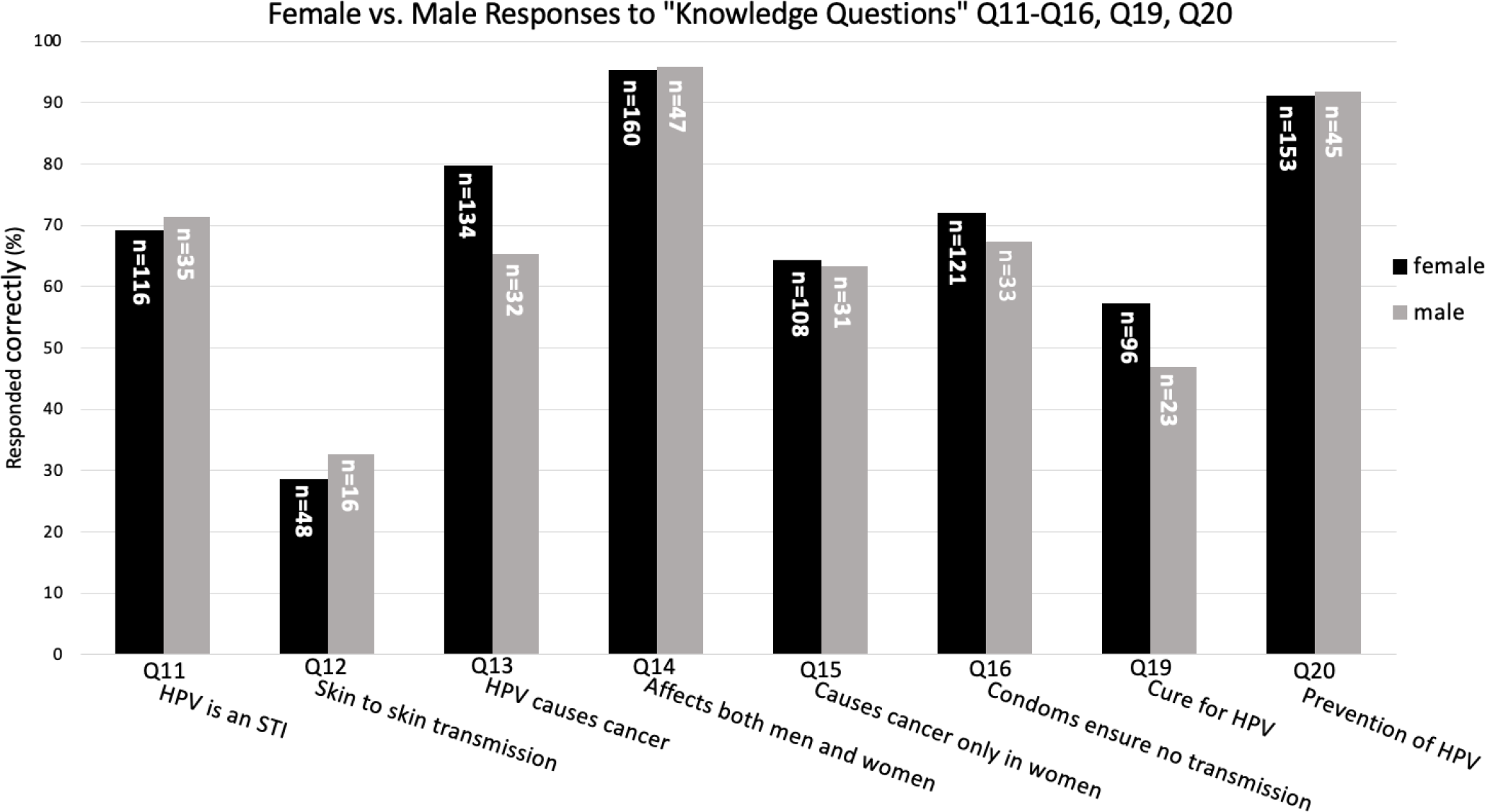
Knowledge about HPV does not play a significant role in influencing vaccination status. The correct response choices to these questions are listed in Table 2.

### Perception on HPV Vaccination Safety

We were also interested in understanding how participants viewed the HPV vaccine itself in terms of safety and protection from infection (**Table 2)**. Females were more likely than males to be aware of the existence of a vaccine for HPV (p=0.004538, χ^2^=8.1369). However, there was no significant difference between female and male response with respect to the knowledge that the newest HPV vaccine, Gardasil-9, approved age range for administration has been expanded. This might not be surprising since this approval for use in women through age 45 is somewhat recent but nevertheless highlights the need to make this information more widespread. Importantly, however, participants (both female and male) who responded that they were aware of the existence of an HPV vaccination were more likely to be vaccinated with at least one dose (responding “yes” to Q23) than expected by chance (p=0.000002134, χ^2^=22.4701).

Females were more likely than males to believe that an individual cannot get HPV from receiving Gardasil 9 (p=0.01266, χ^2^=11.04320). 61.1% of females (n=102) and only 36.7% of males (n=18) responded that an individual cannot get HPV from receiving Gardasil 9. Interestingly, there was no significant difference between biological sex and perception about Gardasil 9 safety and side effects. Greater than 50% of both females and males believed Gardasil 9 vaccination is safe (Q27) and does not have significant side effects (Q26). Importantly, participants (both male and female) who believed Gardasil 9 is safe, effective, without significant side effects, and cannot result in HPV infection were more likely to be vaccinated with at least one dose than expected by chance (p=0.0007145, χ^2^=11.4512). This suggests that information about safety of the vaccine may play an important role in influencing vaccination status.

### Knowledge about HPV vaccination and sexual activity

HPV vaccines are usually administered to adolescents between 11 and 12 years old. At this stage of their lives, it is likely that the decision to vaccinate is made by parents or guardians. One major objection voiced by these parents or guardians is the belief that a vaccine for an STD would make their child more sexually active and/or promiscuous.^28^ We were interested in determining if college aged students shared those beliefs about the HPV vaccine (**Table 2)**. 85.0% of females (n=142) and 75.5% of males (n=37) responded that they would not be more inclined to be sexually active after getting the HPV vaccine. A higher proportion of male participants were unsure if the vaccination would incline them to be sexually active (16.3% of males vs. 4.8% of females). Despite the high proportion of both females and males who believe that vaccination would not impact their sexual activity, females were more likely than expected by chance to believe that the vaccine would not make them inclined to be more sexually active (p=0.02498, χ^2^=7.4384). This result is also in line with other data indicating that HPV vaccine does not increase sexual promiscuity in recipients^29,30,31,32^.

### Approval by family/friends and being vaccinated for HPV

Because HPV is spread, in part, by sexual encounters, some have argued that the vaccine suffers from being sexualized^28^ which suggests that vaccine uptake may be linked to approval from family and friends. We were interested if this kind of ‘approval’, may play a role in the decision to get the HPV vaccine. Interestingly, participants (both male and female) who believed their family/friends would disapprove of the vaccination (responding “yes” to Q30) were less likely to be vaccinated for HPV with at least one dose (responding “no” to Q23) than expected, whereas individuals who did not believe their family/friends would disapprove (responding “no” to Q30) were more likely to be vaccinated (responding “yes” to Q23) than expected (p=.000000008347, χ^2^=37.2028) (**Figure 3)**

**Figure 3:**
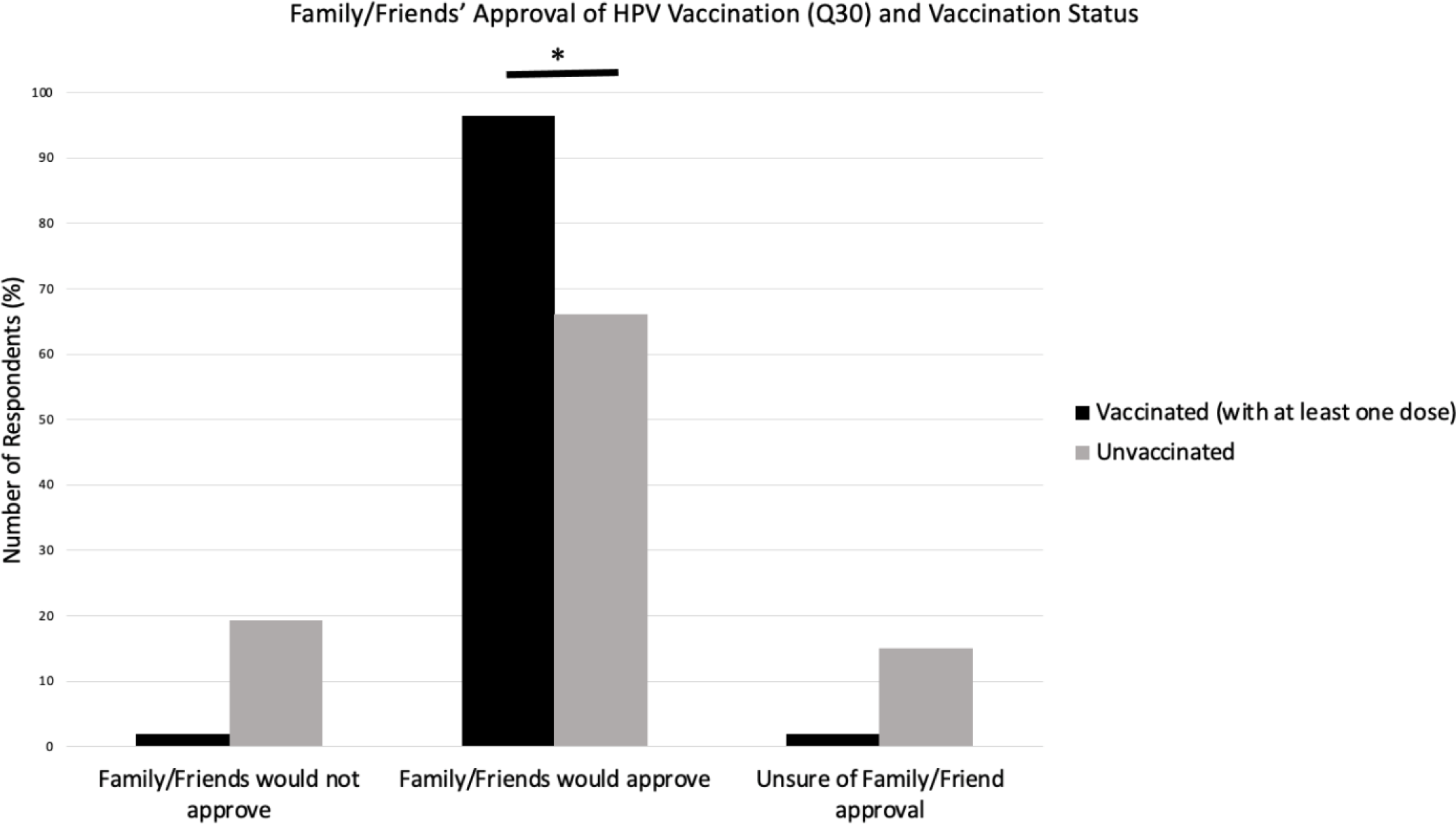
Individuals who feel their family and friends would approve of HPV vaccination are more likely to be vaccinated for HPV. *\*p <0.05*

## Discussion

HPV vaccine uptake remains remarkably low in the United States, with only 71.5% of adolescents being vaccinated with at least one dose in 2019.^10^ College students have the highest prevalence of HPV in the United States along with women visiting STD clinics.^11^ Therefore, we set out to investigate the factors that may influence the decision to be vaccinated in a college aged cohort of students at Villanova University (located in Villanova, PA). We designed a survey to assess knowledge, attitudes, and awareness surrounding HPV infection and HPV vaccination along with self-reported HPV vaccination rates. Our findings suggest that lower vaccination rates for HPV likely involve concerns about vaccine safety and social stigmas associate with HPV, including inclination towards sexual activity and disapproval by friends and family.

### Sex based differences-STD screening

The CDC makes recommendations for STD testing for females and males, although these recommendations differ depending on biological sex, sexual activity, number of sexual partners etc. For example, the CDC recommends that all sexually active people get test for HIV at least once and typically more often depending on sexual activity. Similarly, the CDC recommends that sexually active females under the age of 25, sexually active gay men, or sexually active bisexual men should be screened for gonorrhea and chlamydia annually. Interestingly, the CDC does not recommend annual screenings for STDs for straight men other than where clinically applicable.

Although there are no specific tests for HPV infection, women should be screened for cervical cancers roughly every 3-5 years. The CDC does not make an annual screening recommendation for straight men other than recommending screening for men in high prevalence clinical settings (adolescent clinics, correctional facilities, and STD clinics) or in populations with a high burden of infection (common example is men who have sex with men)^24^. Although women are less likely than men to show symptoms of STDs,^33^ men can still experience asymptomatic STDs.^34^

And since these infections are very efficiently transmitted from males to females,^34^ it is likely these infections will remain prevalent in both men and women. Currently, there is no approved test for HPV in males, and routine screening for anal, penile, and oropharyngeal cancer is not recommended by the CDC.^35^ Therefore, men who develop cancer from HPV infection are usually not diagnosed until the cancer has developed for years or possibly decades.^35^ For this reason especially, it is imperative for boys and men to prevent HPV-related cancers through vaccination with Gardasil-9.

Although we found that females were more likely to be routinely screened for STDs/STIs than males (35.9% vs. 14.3%), we found no significant differences between biological sex and being sexually active, use of safe sex practices, having four or more sexual partners, or a history of diagnosis of an STD/STI. However, our data show that even with the CDC’s annual STD screening recommendation for sexually active women under 25, only 48.0% (n=49) of sexually active females under 25 reported getting routine screening. This statistic is remarkably low and suggests that education about the importance of routine screening combined with information about the benefits of vaccination may provide a means to increase vaccination uptake.

### Sex based differences-Knowledge

HPV is the cause of multiple cancers including anal, cervical, oropharyngeal, penile, vaginal, and vulvar.^2^ In order to better understand why vaccine uptake is low, we wanted to ascertain the general knowledge base about HPV among college aged males and females. Although there were no significant differences between males and females with respect to knowledge about HPV, there was a general lack of knowledge about the virus itself. Only 29.6% (n=64) of males and females correctly responded that HPV can be spread by skin-to-skin contact. Although skin to skin contact is not the most common route of transmission of HPV infection, it is a possible route for transmission and more than half of the study population is misinformed about it. Additionally, 37.5% (n=81) of students surveyed believed that there is a cure for HPV. Despite this being a small subset of the participants, it nevertheless identifies a population of individuals in need of better education about HPV. Better information and education may directly impact their decision to receive the HPV vaccine. Finally, 24.5% (n=53) of males and females were unsure of whether HPV causes cancer only in women (Q15). Although the majority of participants understood that HPV can cause cancer in men and women, this again points out the need for greater education in this area.

The need for improved education is highlighted by the fact that the majority (64.4%, n=139) of participants (both male and female) responded that they felt at a low-risk for getting infected with HPV. This was followed by 17.1% (n=37) responding that they felt at moderate-risk and 13.9% (n=30) at no-risk. Only 4.6% (n=10) participants responded that they felt at a high-risk for getting HPV. Therefore, although HPV is the most commonly transmitted STD/STI in the United States and nearly all sexually active men and women will be infected by some type of HPV in their lifetime^5^, the majority of respondents in our study feel they are at low risk for contracting the virus. Although a conclusive statement cannot be made about the true risk of our study population, 70% (n=151) of participants reported as being currently sexually active, 30.6% (n=66) have had four or more sexual partners, and 66.2% (n=143) report using safe sex practices. It is likely that individuals are more at risk for getting HPV than they feel they are. Interestingly, Holman et al. cite a low perceived risk of HPV infection to be a potential barrier among parents getting their children vaccinated.^36^ This highlights the importance of educating children, adults, and parents alike about the widespread prevalence of HPV and the likelihood of contracting some type of HPV in a person’s lifetime.

Overall, our study found that participants (both male and female) who responded correctly to all eight knowledge questions were not significantly more likely to be vaccinated for HPV (with at least one dose) than expected by chance (p=0.1196, χ^2^=4.1010). This does not preclude the possibility that knowledge is an important factor in the decision to become vaccinated for HPV, but our data suggest that factors beyond knowledge of the infectious disease may play a more significant role in the decision to vaccinate. One explanation for this could be that the majority of individuals become vaccinated for HPV through the choice of their parent(s)/guardian(s) so the child’s individual knowledge may not correlate with vaccination status. Proper education is still vitally important because these same individuals who were vaccinated as children will likely be making the decision whether or not to vaccinate their own children for HPV. While our data does not suggest a correlation between an individual’s knowledge and their own HPV vaccination status, but correct knowledge is critically important surrounding the topic of HPV and should be increased in the aforementioned areas.

### Safety

Individuals who believed Gardasil 9 is safe, effective, without significant side effects, and cannot result in HPV infection were significantly more likely to be vaccinated in at least one dose than participants who believed Gardasil 9 is unsafe. Scientific literature proves the safety and efficacy of Gardasil 9, but a divide exists between these facts and the public’s understanding of them.

### Inclination towards sexual activity

There is a perception held by some parents that getting their child vaccinated for HPV will incline the child to become sexually active. This is commonly linked to a feeling by such parents that getting their child vaccinated at the recommended age of 11-12 years old sends a message to the child that it is okay or even encouraged to be sexually active following HPV vaccination.^37^ Our findings add evidence to dispel this notion. We found that 85.0% of females (n=142) and 75.5% of males (n=37) responded that they will not be more inclined to be sexually active after getting the HPV vaccine. This unsupported perception has the potential to be a detriment to the health of the child through not vaccinating him or her against HPV or waiting until an older age to vaccinate which is inadvisable. A physician’s recommendation for the HPV vaccine has been shown to promote uptake and acceptance of HPV vaccination.^36^ Therefore, physicians can be an important target to promote HPV vaccination and alleviate concerns about its link to sexual activity which is widely disproven^29–32^.

### Disapproval

Parental disapproval of HPV vaccination can be a detriment to HPV vaccination not only in children but also in young adults. This survey found that individuals who believe their family or friends would disapprove of them getting the HPV vaccination were statistically significantly less likely to be vaccinated than individuals who did not believe their family or friends would disapprove. It is imperative that repeated educational conversations occur between parents, children, and healthcare providers about HPV vaccination to eradicate its stigma with sexual promiscuity or inclination. If individuals feel supported by their family and friends to become vaccinated for HPV, the evidence from our study supports that they are more likely to do so. Stigma plays a major role in preventing HPV vaccination, and it is vital that this stigma is replaced by perceptions that align with statistical facts. The data in our survey shows that HPV vaccination does not incline individuals towards sexual activity.

Overall, our study suggests that education about HPV and HPV vaccination may have important impacts on vaccination rates. Since the recommended age range for HPV vaccination is 11-12 years of age, clinicians should discuss the virus and vaccine with both parents *and* children and, if possible, dispel the myths that surround HPV vaccination. Our study also suggests that even when children are not vaccinated, or not fully vaccinated, against HPV their continues to be opportunities to education these individuals as they move through the health care system, particularly in early adulthood.

## Data Availability

Aggregated data available upon request. No individualized responses will be shared to protect anonymity of respondents.

## Acknowledgements

We would like to thank Dr. Carol Weingarten for her guidance and suggestions during and after the data collection process, and Dr. Carla NavarezDiaz for helpful comments and discussion about the statistical tests used to analyze the collected data. We would also like to thank Dr. Carol Weingarten, Dr. John Olson, the Villanova Honors Department, and the Office of Health Professions Advising for help in disseminating the survey.

## Author Contributions

JAG conceptualized the study, collected responses, and performed statistical analyses. JAG and JDC designed the survey and the plans for dissemination and wrote the paper. No competing interests are declared.

